# Evaluation of two easy-to-implement digital breathing interventions in the context of daily stress levels in a series of N-of-1 trials: results from the Anti-Stress Intervention Among Physicians (ASIP) Study

**DOI:** 10.1101/2025.04.09.25325513

**Authors:** Valentin Max Vetter, Tobias Kurth, Stefan Konigorski

## Abstract

Physicians face intense work-related stress, which can harm their health, increase the risk of medical errors, lower healthcare quality, and increase costs within the healthcare system. In this 4-week intervention study, individual-level and population-level effects of two short and easy-to-perform breathing exercises designed to reduce stress are evaluated among 76 physicians in residency in Germany in a series of N-of-1 trials. Levels of stress and levels of stress expected for the following day were assessed electronically every day via the StudyU App. Intervention effects were estimated using Bayesian linear regression models and were overall small on the population level but they showed large heterogeneity between individuals with strong effects for selected individuals. Twenty-seven participants benefitted from the anti-stress exercises. Three (Mindfulness Breathing) and seven participants (box breathing) had a ≥70% probability for a daily stress reduction of ≥0.5 points and thereby fulfilled our responder criteria. On average, 4.5 months after completion of the individual N-of-1 trials, a follow-up survey among all participants was conducted to assess sustainability of the interventions. Although general implementation of these anti-stress exercises cannot be recommended, they can be highly effective for selected individuals and potentially beneficial in specific subgroups.

## Introduction

Physicians are regularly exposed to work-related and emotional stressors^1,2^, which, together with inadequate self-care^3^, are responsible for the frequently reported high stress burden in this group (reviewed in ref.^2^). The rate of burnout and emotional distress among physicians was shown to be between 23% and 76%^4–11^ and 22% of German physicians in residency reported to have taken medication because of work-related stress^12^. Differences in burnout rates between specialties were found^7^ and the stress burden was higher in younger physicians and physicians in residency^2,5,13^.

Exposure to acute stress activates two main physiological systems. The sympatho-adrenal medullary (SAM) system releases catecholamines, which affect various physiological responses, including heart rate, blood pressure, and the pulmonary system, preparing the body for a ‘fight or flight’ response when threatened^14,15^. The hypothalamo-pituitary adrenal (HPA) axis causes the release of glucocorticoids which have anti-inflammatory effects, promote gluconeogenesis, and modulate various metabolic processes^15–17^. A frequent or prolonged activation of these systems increases the risk for diseases^18,19^ mostly through their immune-modulatory effects^20,21^. Additionally, high levels of stress are associated with unhealthy behaviors such as sleep deprivation and decreased exercising^16^, increased nicotine and alcohol consumption^22,23^, substance abuse^24^, and impulsive decision making^25^. The association of psychological stress, disease and other adverse health outcomes for the individual experiencing stress is well-established^16,19,26–28^. However, in physicians, high levels of stress and burnout were shown to have effects exceeding the individual level as they were associated with increased risk for deviation from medical standards and major medical errors, poorer doctor-patient relationships, and overall lower quality in patient care^5,29–38^. Therefore, reducing stress levels among physicians would increase the physical and mental health of the medical workforce and could potentially improve patient care^39^. The reduction of stress in physicians is also of economic interest^40^. High levels of stress were shown to be associated with high turnover rates, burnout, early retirement, and lower productivity rates among physicians^35,41–44^, which in turn substantially increases costs within the healthcare system^2,40^. For this reason, the Anti-Stress Intervention Among Physicians (ASIP) study evaluates two easy-to-learn breathing interventions that can be flexibly performed and, do not require special training or additional tools and are therefore easily implemented into everyday routine. The first intervention, a guided 8-minute mindfulness and breathing exercise^45,46^, includes stretching and simple upper body movements. The second intervention, Box Breathing (also known as “tactical breathing”), was shown to decrease the heart rate and is used by military and law enforcement to cope with stressful situations^47–49^. Beneficial effects of breathing exercises^46,50,51^ and mindfulness interventions^39,45,52,53^ on stress, anxiety, and well-being were shown before in some studies, but other studies did not find a statistically significant effect of these interventions^45,51,54,55^. A large degree of heterogeneity of the individual intervention effects is expected^56^ and was previously reported for mindfulness based anti-stress interventions^57^. To assess individual-level effects, populations-level effects, and between-participant differences in the intervention effect, we conducted a large series of N-of-1 trials^58^. Each participant went through a randomly allocated pre-specified 4-week sequence of intervention and control phases and reported their stress level on each day. After trial completion, the individual-level intervention effects were assessed by comparing the daily participant reported outcomes (PRO) between phases. The population-level intervention effect was estimated by aggregating the data of the individual N-of-1 trials. This study design is considered to be the gold standard when dealing with strong effect heterogeneity^58^ and allows for individual feedback on the observed effects to each participant. On average 4.5 months after completion of the individual N-of-1 trial, a longitudinal follow-up was conducted to assess sustainability of the evaluated interventions. A carefully developed study design guaranteed full anonymity of all collected data while allowing horizontal cross-platform data linkage as well as longitudinal assessments.

In this study, we report on the individual-level and population-level effect of two anti-stress interventions, Mindfulness Breathing and Box Breathing, on the average daily level of stress and the average level of stress expected for the following day in 76 physicians in residency in Germany. We hypothesized that performing an anti-stress intervention reduces the daily stress level and the level of stress expected for the following day^59^. The ASIP study was conducted in accordance with the previously published study protocol^59^.

## Results

### Study Participants

Of the 8,032 confirmed recipients of the study invitation, 359 participants indicated interest in participation and signed up to receive the study information and consent form. Of these participants, 125 completed the questionnaire. Thirty-three chose the Mindfulness Intervention (461 PROs documented over all trials), while 43 opted for the Box Breathing intervention (662 PROs documented over all trials, Figure 1). Each N-of-1 trial consisted of a randomly allocated alternating sequence of intervention (A) and control (B) phases (ABAB or BABA), where each intervention phase lasted 1 week for a total trial length of 4 weeks, and participants were asked to document the PROs daily. Participants were guided through their individual trial and reminded to perform the intervention by the StudyU App installed on their smartphone. Across interventions, 49.4% of all PROs were documented during the intervention periods.

**Figure 1:**
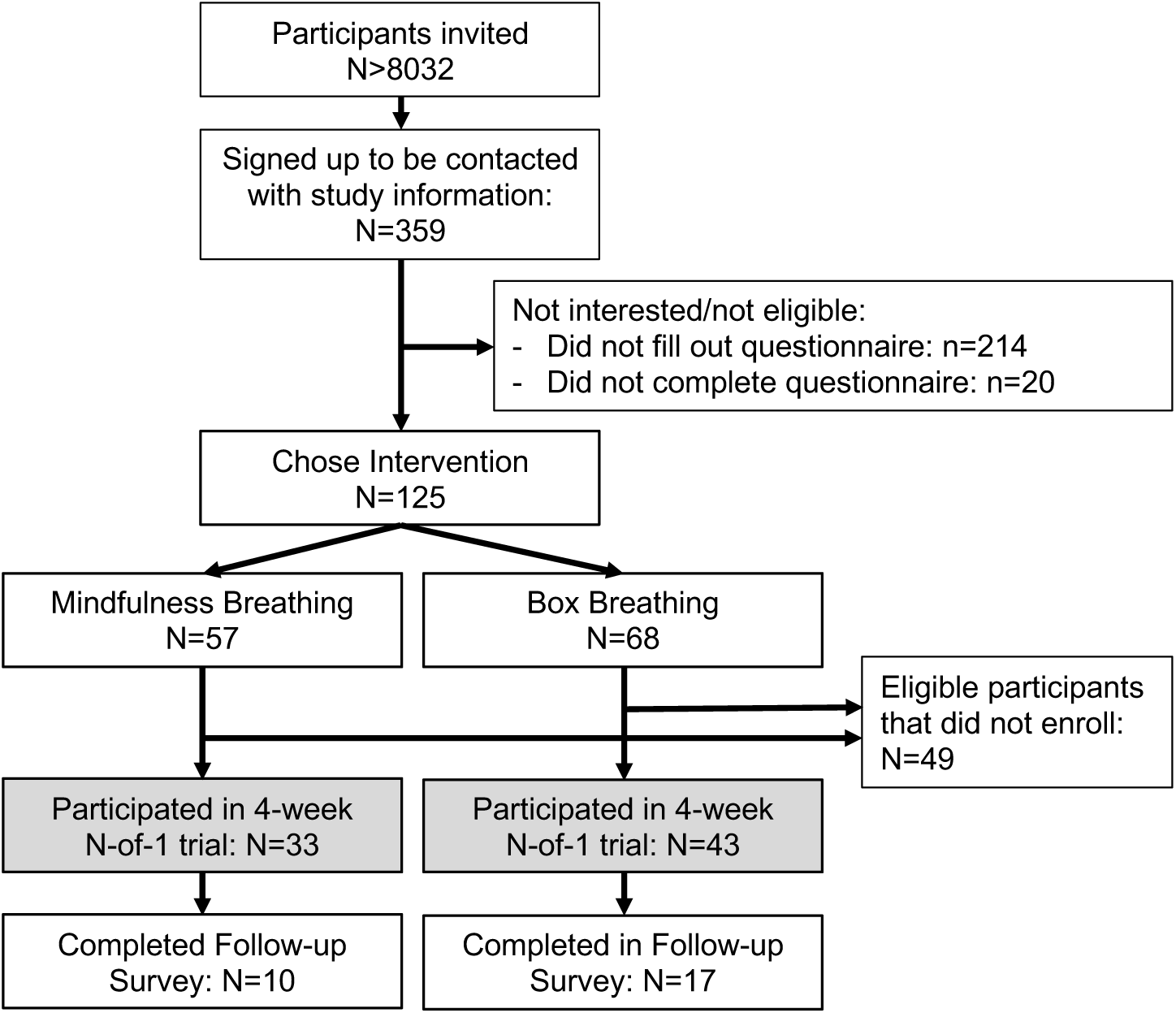
Chart of participants flow in the ASIP study. After recruitment, participants were screened for eligibility and asked to fill out a baseline questionnaire, at which end they were asked to choose which intervention they would like to perform during their individual 4-week N-of-1 trial. On average, 4.5 months after their individual N-of-1 trial ended, participants were asked to fill out a follow-up survey.

Participants were, on average, 35 years old, and 80% were women. Their overall self-rated health was good or very good (88%), and 65.8% of the participants reported drinking alcohol less than once per week. A large majority reported having never smoked (76.3%). Participating physicians in residency were, on average, in their fourth year of clinical training with, on average, 35 contractually agreed working hours per week (Table 1). Participants generally showed high values in the psychological questionnaires aimed at quantifying individual chronic stress, burnout symptoms, work-related privacy conflicts, over-commitment at work, and a low effort reward ratio. Life satisfaction was in a range comparable to the general public (Table 1). Chronic stress levels, assessed using Cohen’s 10-item Perceived Stress Scale (PSS), were correlated with patient-reported outcomes (PROs) of stress assessed at baseline. Specifically, chronic stress showed a moderate correlation with daily stress levels (Pearson’s r = 0.5, 95% confidence interval: 0.3 to 0.6) and a weak correlation with anticipated stress levels for the following day (Pearson’s r = 0.2, confidence interval: 0.0 to 0.4). Lower average stress levels were documented during the weekend (Supplementary Figures 1 and 2).

**Table 1:**
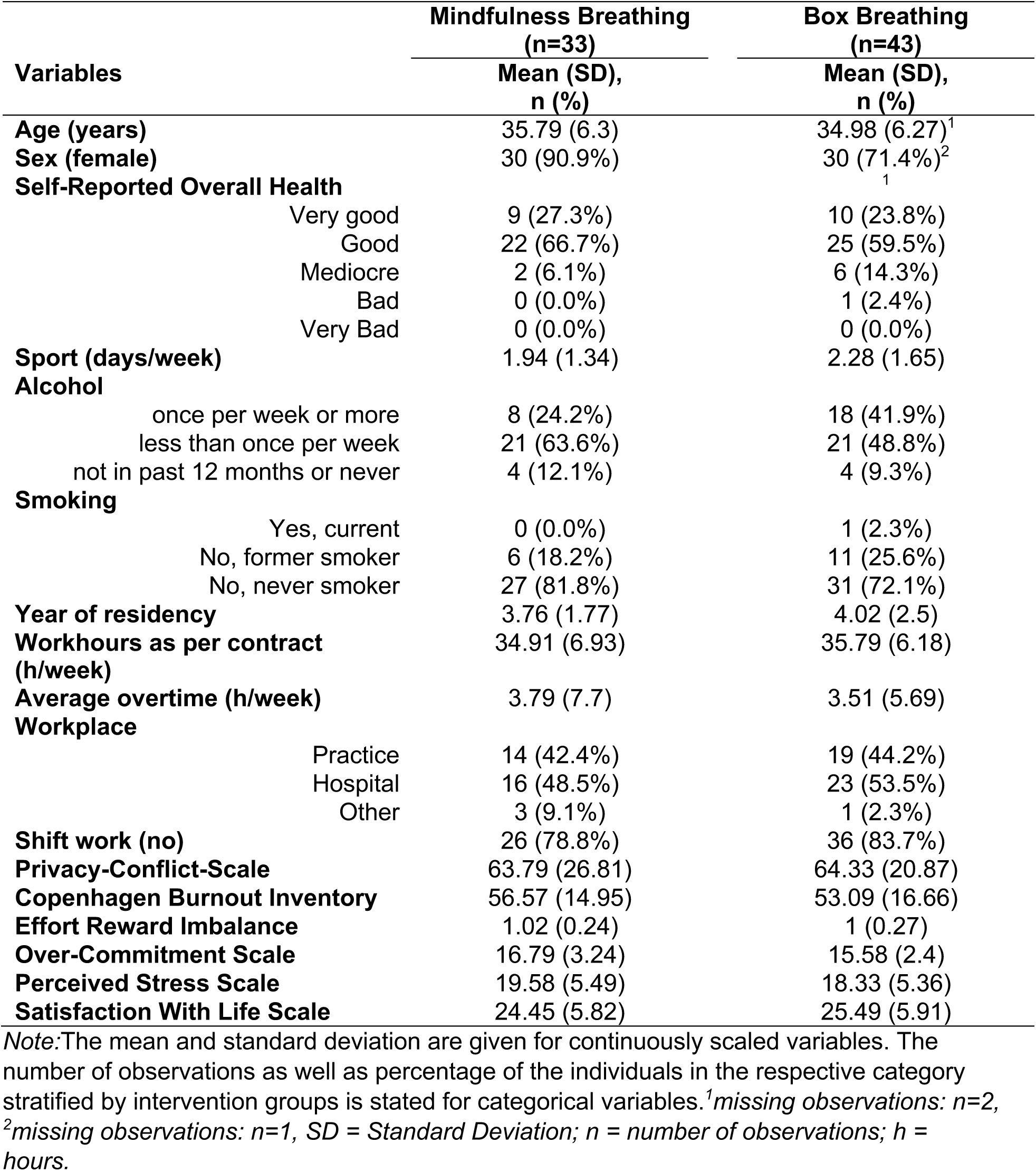
Baseline data on participants was assessed via questionnaire prior to the start of the individual N-of-1 trials.

### Individual-level analyses of each N-of-1 trial

In a first step, the individual intervention effect was estimated separately for each participant using Bayesian linear regression models with an autoregressive error structure (AR1). To evaluate the individual level intervention effect, we reported the estimated mean of the posterior distribution, its 95% credible interval, and the estimated probability of reaching a stress reduction of at least 0.5 points, which was defined to be clinically relevant before data assessment. Participants with a 70% or higher probability of achieving this effect were classified as responders.

With respect to daily stress levels, substantial heterogeneity in the posterior mean estimate of the intervention effect was observed. It ranged between −1.4 and 3.1 among participants who chose Mindfulness Breathing (Figure 2 A, Supplementary Table 1) and between −3.0 and 1.5 in the group that chose Box Breathing (Figure 2B, Supplementary Table 2). In the group that performed Mindfulness Breathing, 14 participants showed a posterior mean below zero (i.e., showed decreased stress levels during the intervention phases), and three were classified as responders to decreasing stress levels during the intervention phases (Figure 2 C). Among participants performing Box Breathing, 17 participants had a negative mean posterior, of which seven fulfilled the responder criteria (Figure 2 D). A negative effect estimate indicated a reduction in daily stress levels in points on a 0 to 10 scale.

**Figure 2:**
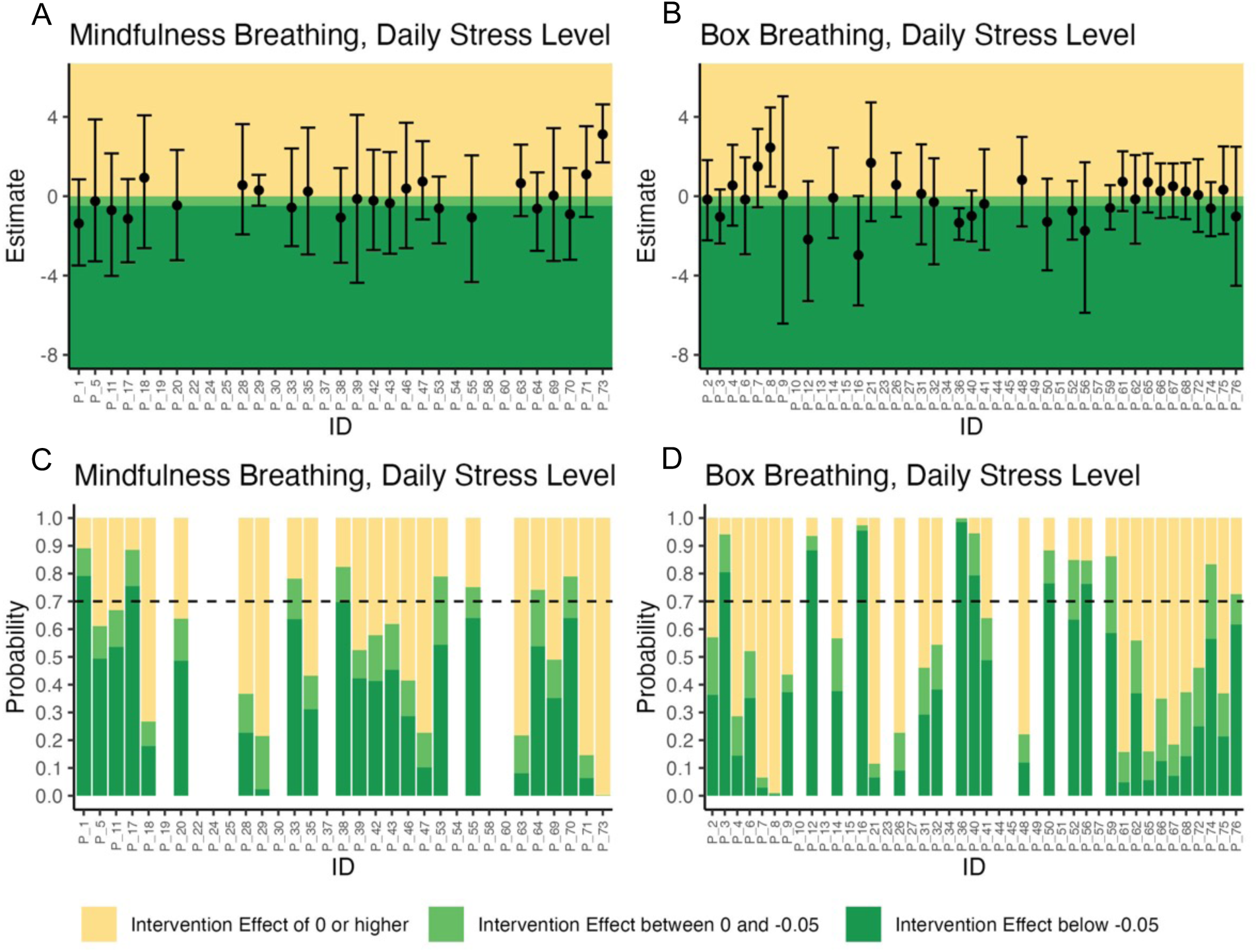
Results of the 76 individual N-of-1 trials (shown on x-axis) with respect to daily stress levels. A and B: Estimates of posterior mean and 95 % credible interval of the difference in daily stress levels between intervention and control periods. B and C: Estimated posterior probability of a reduction of daily stress levels and reaching a clinically relevant stress reduction of 0.5 points or more. If the probability of stress reduction was 70% or higher, the participant was classified as a responder).

Similarly, heterogeneous results were found for the expected stress levels on the next day, with mean posteriors ranging between −1.7 and 2.3 (Mindfulness Breathing, Figure 3 A, Supplementary Table 3) and −1.6 and 2.0 (Box Breathing, Figure 3 B, Supplementary Table 4). Among participants performing Mindfulness Breathing, 13 had posterior means below zero, and two were identified as responders (Figure 3 C). Similarly, 16 of the study participants in the Box Breathing arm had mean posteriors below zero, and three met the responder criteria (Figure 3 D).

**Figure 3:**
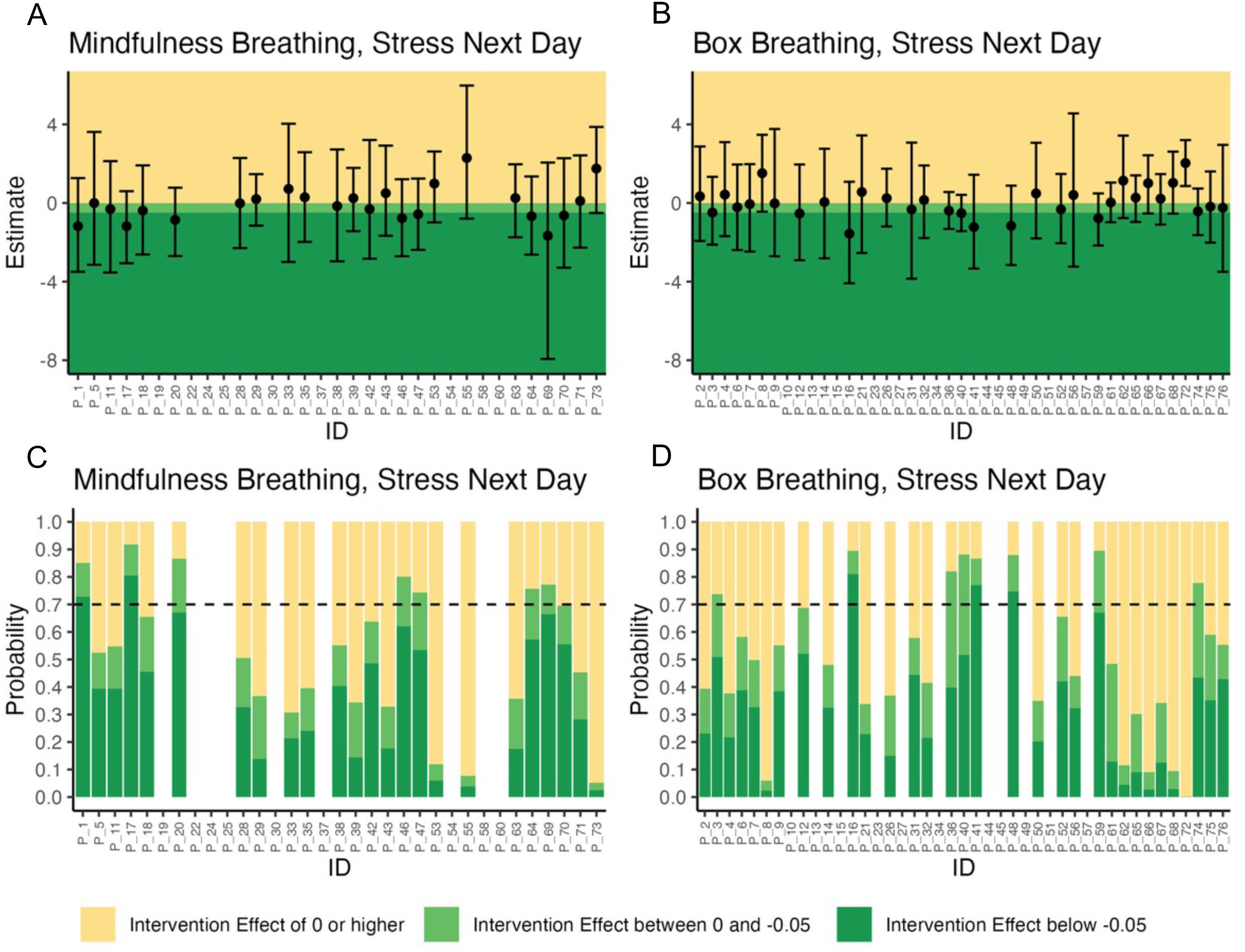
Results of the 76 individual N-of-1 trials with respect to levels of stress expected for the following day. A and B: Estimates of posterior mean and 95 % credible interval of difference in expected levels of stress of the next day between intervention and control periods. B and C: Estimated posterior probability of a reduction of expected level of stress on the next day and reaching a clinically relevant stress reduction of 0.5 points or more. If the probability of a clinically relevant stress reduction was 70% or higher, the participant was classified as a responder.

The individual level time series data of reported stress levels are shown in Supplementary Figures 1 and 2.

### Population-level analyses of each anti-stress intervention

On the population level, we estimated the average effect of each intervention among the population that decided to participate in the respective intervention on the two primary outcomes (daily stress level and level of stress expected for the following day). Bayesian multilevel linear regression models with AR1 error structure were used to estimate the mean posterior, the 95% credible interval, and the probability of reaching a clinically relevant effect. On average, among participants that chose to perform Mindfulness Breathing, the intervention reduced the daily stress level by 0.18 points (95% credible interval: −0.62 to 0.26). The probability of reducing the daily stress levels by performing the intervention was 79.3%, and achieving a clinically relevant reduction of at least 0.5 points was 7.7% (Figure 4). Among participants doing Box Breathing, an average reduction of daily stress levels by 0.07 points (95% credible interval: −0.44 to 0.32) was observed. The probability of reducing the average daily stress level using Box Breathing was 64.0%, and reaching a clinically relevant reduction was 1.3% (Table 2, Figure 4).

**Figure 4:**
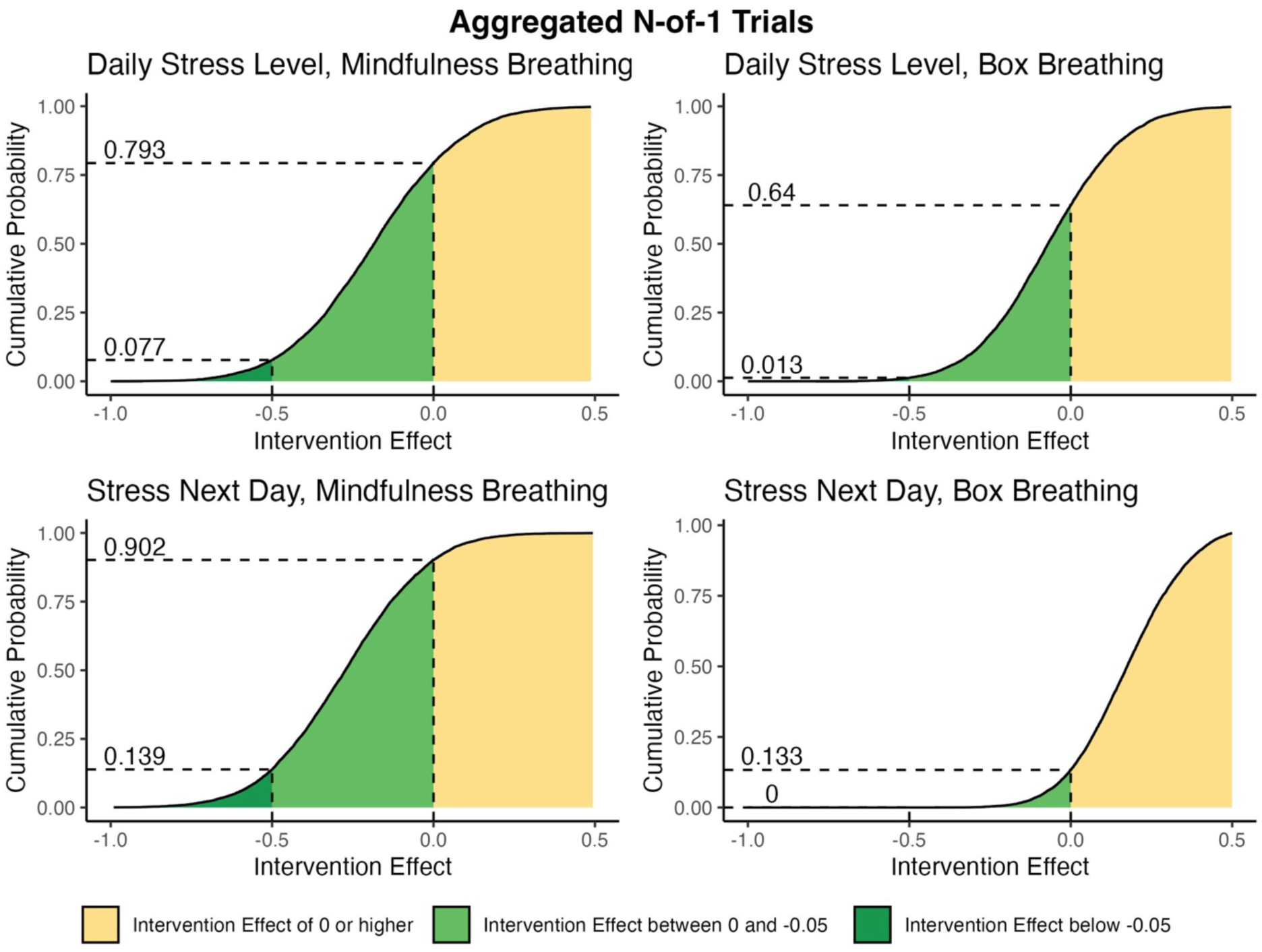
Estimated cumulative probability for the intervention effect of the anti-stress interventions on the daily stress level and level of stress expected for the next day to be higher than a given value (on the x-axis). Probabilities for a stress level reduction (difference <0) and for a clinically relevant effect (difference < −0.5) are marked with dotted lines.

**Table 2:**
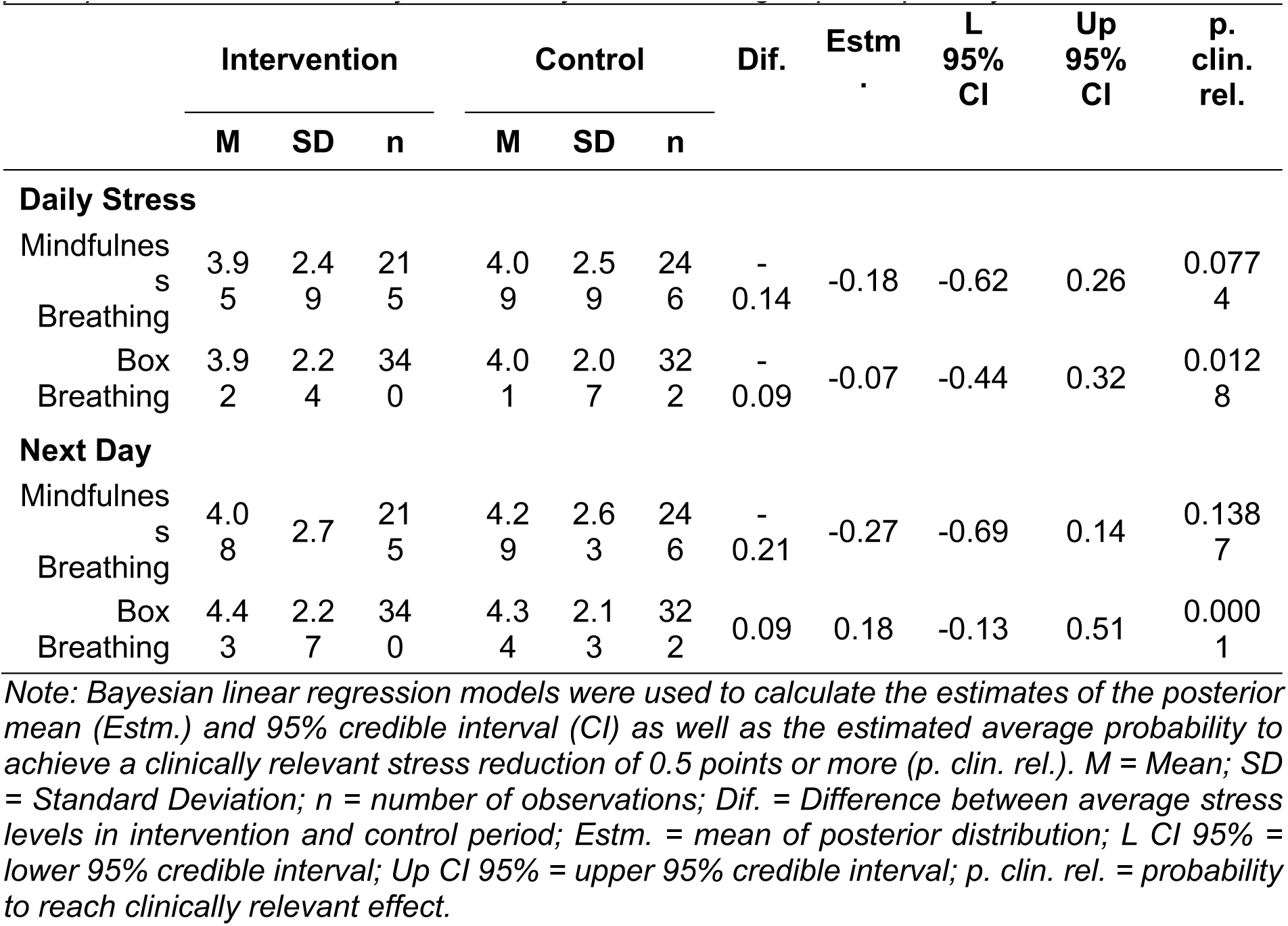
Table of mean (M), standard deviation (SD), number of documented PROs (n), and mean difference in outcome variables between intervention and control periods (Dif.) in 76 participants of the ASIP study stratified by intervention group and primary outcome.

For the level of stress expected for the next day, Mindfulness Breathing resulted in a reduction of 0.27 points (95% credible range: −0.69 to 0.14). The probability of reducing the expected stress level was 90.2%, and achieving a clinically relevant reduction was 13.9%. Among participants using Box Breathing, the intervention increased 0.18 points (95% credible interval: −0.13 to 0.51) of stress expected for the next day. Therefore, the probability of achieving a clinically relevant reduction in stress was <0.01% (Table 2).

Secondary analyses: effect measure modifiers and average intervention effect of intervention-availability across study arms To exploratively analyze potential effect measure modifiers, subgroup analyses stratified by sex and chronic stress level at baseline were performed. In the subgroup of women, we observed effect sizes similar to the complete dataset. However, the estimated intervention effect of Mindfulness Breathing was less strong. In the subgroup of men, performing the Mindfulness Breathing intervention reduced daily stress levels by 1.25 points (95% credible interval: −4.32 to 2.40), and a clinically relevant reduction of stress by 0.5 points had a probability of 75%. Similarly, performing this intervention reduced the level of stress expected for the following day on average by 0.57 points (95% credible interval: −6.77 to 3.39), and the probability of reaching a clinically relevant stress reduction was 48.4% among men. Interestingly, Box Breathing was estimated to result, on average, in an increase of daily stress among male participants (mean posterior: 0.1, 95% credible interval: −0.69 to 0.95) as well as of level of stress expected for the next day (mean posterior: 0.24, 95% credible interval: −0.44 to 0.96, Supplementary Table 5).

In the subgroup of participants with low chronic stress at baseline (<50^th^ percentile of Cohen’s PSS), Mindfulness Breathing did not lead to a reduction in daily stress levels (posterior mean: −0.01, 95% credible interval: −0.71 to 0.71) or stress levels expected for the next day (posterior mean: −0.05, 95% credible interval: −0.67 to 0.59). However, a reduction of 0.32 points in the stress level on each day (95% credible interval: −0.97 to 0.31) was observable among participants performing Box Breathing and 0.17 points in the level of stress expected for the following day (95% credible interval: −0.65 to 0.36). In the group of participants with high levels of chronic stress, lower daily stress levels (mean posterior: −0.27, 95% credible interval: −0.95 to 0.40), as well as lower levels of expected stress (mean posterior: −0.29, 95% credible interval: −0.97 to 0.38), were found among participants evaluating Mindfulness Breathing during the interventions periods. However, among participants evaluating Box Breathing, higher stress levels in this group (daily stress: mean posterior: 0.13, 95% credible interval: - 0,35 to 0.64; expected stress next day: mean posterior: 0.38, 95% credible interval: −0.06 to 0.83, Supplementary Table 6) were documented after performing the intervention.

Finally, by aggregating both groups, we estimated the average effect of performing an anti-stress intervention among participants using the intervention of their choice. No intervention effect was observable when analyzing the aggregated data from both study arms together (Supplementary Table 7).

### Follow-up

A follow-up questionnaire was sent to all participants on average 4.5 months after the individual N-of-1 trials ended. Successful matching of a completed follow-up questionnaire with baseline data was possible for 10 (Mindfulness Breathing) and 17 participants (Box Breathing). Across interventions, 58% of participants felt that they benefitted from performing the intervention, and 42% planned to use it in the future. Box breathing was recommended to others by 47,1%. However, only 30% of participant evaluating Mindfulness Breathing recommended the intervention to others. While 40% of the participants from the Mindfulness Breathing group performed the intervention after completion of the N-of-1 trial, more than 82% of participants from the Box Breathing group report using the intervention in their daily life (Table 3). Of all participants who filled out the follow-up questionnaire, one reported trouble breathing and dizziness after performing Box Breathing.

**Table 3:** Descriptive statistics of the response collected as part of the follow-up questionnaire which was sent out to all participants on average 4.5 months after completion of the individual N-of-1 trials.

|  | Mindfulness<br>Breathing<br>n(%) | Box<br>Breathing<br>n(%) |
| --- | --- | --- |
| **The intervention has reduced my stress level.** |  |  |
| Disagree | 0 (0.0) | 1 (6.2) |
| Somewhat Disagree | 4 (40.0) | 6 (37.5) |
| Somewhat Agree | 6 (60.0) | 8 (50.0) |
| Agree | 0 (0.0) | 1 (6.2) |
| **I plan to use the intervention allocated to me in the future.** |  |  |
| Disagree | 1 (10.0) | 1 (6.2) |
| Somewhat Disagree | 6 (60.0) | 7 (43.8) |
| Somewhat Agree | 3 (30.0) | 5 (31.2) |
| Agree | 0 (0.0) | 3 (18.8) |
| **Have you recommended the intervention to someone else?** |  |  |
| No | 7 (70.0) | 9 (52.9) |
| Yes, to a colleague | 2 (20.0) | 2 (11.8) |
| Yes, to a friend | 1 (10.0) | 1 (5.9) |
| Yes, to my family | 0 (0.0) | 1 (5.9) |
| Yes, to patient | 0 (0.0) | 4 (23.5) |
| **Do you use the intervention in your everyday life?** |  |  |
| Yes, daily or almost daily | 0 (0.0) | 1 (5.9) |
| Yes, on 1-6 days per week | 1 (10.0) | 4 (23.5) |
| Yes, less than once per week | 0 (0.0) | 0 (0.0) |
| Yes, once a month or less | 3 (30.0) | 9 (52.9) |
| No, never | 6 (60.0) | 3 (17.6) |

Note: n = number of observations.

## Discussion

In this study, we evaluated the effect of two short anti-stress interventions on the daily stress level and on the stress level expected for the following day in 76 physicians in residency in Germany. Due to an expected high heterogeneity in intervention effects between individuals, a series of N-of-1 trials was conducted to estimate the individual-level and the population-level effect separately for each intervention using Bayesian linear regression models. Large between-individual differences were observable for both anti-stress interventions, with mean posterior estimates ranging between −3 and 2.3. A total of 27 participants experienced benefits from performing one of the anti-stress exercises. To further examine differences in the intervention effect, sex- and stress-stratified subgroup analyses were conducted. On the population level, a small reduction of the daily stress level was found for both Mindfulness Breathing and Box Breathing which is in line with our hypothesis. While the mindfulness intervention also reduced the expected stress levels for the following day, participants in the Box Breathing group, contrary to expectations, exhibited a slight increase in stress after performing the intervention. Our exploratory secondary analyses observed differences in the intervention effects between sex- and stress-stratified subgroups. In men, participants performing Mindfulness Breathing showed reduced stress levels, while among those performing Box Breathing, an increase in the perceived level of stress was observable. In the subgroup of women, no substantial differences to the results from the complete dataset were found. Participants with low levels of chronic stress benefitted from Box Breathing, but no effect of Mindfulness Breathing was found. Among participants with high levels of chronic stress who performed Mindfulness Breathing a reduction of stress levels during the intervention periods was documented. These results are in line with previous findings, which also reported stronger effects in participants with high baseline stress^60^. However, in highly stressed participants of the Box Breathing group, a slight elevation of stress perception was observed during the intervention phases compared to the control phases.

The large heterogeneity of the intervention effects suggests that the performance of breathing interventions can be highly effective for selected individuals but might not lead to a strong average stress reduction in larger groups. This was also observable in the exploratory subgroup analyses, which showed the largest intervention effects for Mindfulness Breathing among men and participants with high baseline stress.

The observed overall reduction in daily stress levels in this study due to the performance of anti-stress exercises is in line with previously published intervention studies conducted among physicians^61–64^. However, the population level effect sizes found in this study are smaller compared to previous results. This could potentially be due to the short overall study duration, shorter and less intensive anti-stress interventions, and differences in the analyzed study population^61–64^. Smaller effects of shorter interventions compared to longer interventions or structured programs (meta-analyzed in ref.^65^) were reported before, and a dose response relationship between intervention length and stress reduction was suggested^66^. In this study, we aimed to compensate for the short duration and low dose of the interventions by instructing participants to perform the anti-stress intervention daily, which has been noted to increase the effect of anti-stress interventions^66^.

There are a number of limitations to this study. The interventions were designed to be self-administrable and to be performed at the time of the participants convenience. Therefore, differences in the time between intervention and documentation of PROs as well as the time of day at which the intervention is performed are expected. Also, a large variance in the documented daily levels of stress were found which might limit our ability to detect intervention effects. In future studies, a daily assessment of systematic confounders could help to reduce the variance in the outcome measures.

Strengths of this study include the N-of-1 study design which allows for the estimation of individual-as well as population-level effects. The large effect heterogeneity found in our study underlines the importance of using study designs which are able to analyze individual-level effects as well as between-person differences when evaluating anti-stress interventions. Another strength of the study is its large sample size of 76 participants, which is more than twice the number of participants required (n=34) according to our sample size calculation^59^.

In summary, performing Mindfulness Breathing and Box Breathing results in a small reduction of daily stress levels. Large differences in the intervention effect were shown between participants and in secondary subgroup analyses. The most notable reduction in daily stress was observed among men and participants with high levels of chronic stress at baseline who practiced Mindfulness Breathing. Among participants evaluating Box Breathing, a reduction in stress levels during the intervention periods was observed for participants with low baseline stress, but a small increase in stress levels among men or participants with high baseline stress was found compared to control periods.

Mindfulness Breathing. Therefore, while our findings do not support the general application of the evaluated anti-stress interventions among physicians in residency, carefully selected individuals may benefit substantially from performing one of the two anti-stress interventions. Additionally, our findings emphasize the importance of using study designs able to investigate effect heterogeneity when analyzing anti-stress interventions.

## Methods

### Trial design

The Anti-Stress Intervention Among Physicians Study (ASIP) aims at the evaluation of two anti-stress interventions among healthy physicians in training in Germany. After screening for eligibility, participants were asked to fill out a baseline questionnaire and indicate their preference for Mindfulness Breathing or Box Breathing. The control condition was everyday life. Subsequently, participants were referred to the StudyU App^67^ and provided with an individual randomized sequence of two pairs of one-week intervention (A) and control (B) periods resulting in a total trial duration of 4-weeks and two possible intervention-control-sequences (ABAB or BABA). In the StudyU App, participants were electronically guided through the 4-week study period and were provided with the digital anti-stress interventions. Three months after completing the collection of data from the individual N-of-1 trials, a follow-up questionnaire was sent out to all participants via email.

### Participants

The study was conducted in Germany among healthy physicians in training. Inclusion criteria were weekly working time in a clinical position of at least 9 hours, regular access to a smartphone on which the StudyU App can be installed, and informed consent. Participants were excluded if they were <18 years of age, had already completed their specialist training, had no clinical activity during the study period (e.g. vacation, research activity, etc.), participated in another intervention study during the study period, did not speak German, performed yoga more than 4 times a month, meditated or performed breathing exercises on average more than 4 days per month, were confirmed or suspected to be pregnant or had a psychiatric disorder, cardiovascular disease, respiratory, pulmonary or neurological disease. Furthermore, participants were excluded from the study if they had a planned surgery within the next 6 months, reported substance abuse (for example, alcohol, drugs, or other), or a physician recommended (or self-assessment) not to perform mindfulness or breathing exercises. Due to data protection reasons, employees of the Charité - Universitätsmedizin Berlin were not eligible for study participation.

Recruitment of participants took place between April 15, 2024, and May 31, 2024, and was done via email, posting of study invitations in local messenger groups and on websites of institutes of university hospitals, as well as displaying the study invitation on seminar days for physicians in training. Interested participants were asked to provide their email addresses through a contact form and were subsequently sent the study information, the consent form, and a link to the baseline questionnaire. Additionally, participants were provided with contact information (email and phone number) of the study PI to discuss questions and obtain further information.

All participants provided informed consent. The study was conducted in accordance with the Declaration of Helsinki and approved by the Ethics Committee of the Charité-Universitätsmedizin Berlin - approval number EA4/260/23. The trial is registered at ClinicalTrials.gov under trial ID NCT06368791 (first registered April 11, 2024). No changes were made to the originally published study protocol^59^.

### Procedure

Data collection took place between April 15, 2024, and June 30, 2024. The carefully developed study design guaranteed anonymity of all collected data while maintaining cross-sectional and longitudinal data linkage across platforms used for data collection. Before beginning with the individual N-of-1 trials, participants were asked to fill out a baseline questionnaire at the end of which they were given the opportunity to choose which intervention they wanted to do as part of the study. Participants then received an individual linkage code to get access to the study in the StudyU App^67^. This mobile app for both iOS and Android phones was used to guide each participant through their individual trial, deliver the anti-stress interventions, and document the daily PROs. Participants were informed about their individual intervention-control-sequence and were reminded by push notifications to perform the intervention (during intervention periods) and to document the PROs (during intervention and control periods). Anonymous data collected through the REDCap questionnaire and the StudyU App is publicly available (see below). Furthermore, the setup of the ASIP study, including the detailed N-of-1 procedure, as well as the respective interventions is made publicly available for full transparency and easy replication within the StudyU App in the StudyU Designer (https://designer.studyu.health).

### Interventions

In this trial, two anti-stress interventions were tested independently. The first, Mindfulness Breathing consisted of an 8-minute guided mindfulness and breathing exercise. This intervention was delivered as an audio file through the StudyU App. Participants were asked to find a comfortable seated position in a quiet and low irritation environment. They were then instructed to perform a number of breathing exercises alongside simple body movements and stretching exercises. During the intervention, participants were advised to pay attention to conscious breathing, follow the flow of air through their body and to focus on the feeling of calmness as a result of their natural breathing cycle. The second intervention, Box Breathing, was a 6-minute structured breathing exercise presented as a video file. This video was seamlessly integrated and played within the StudyU App. Participants were instructed to breathe in for four seconds, hold their breath for four seconds, and breathe out for four seconds. After holding their breath for a further four seconds, the next breathing cycle began with four seconds of breathing in. Participants were provided with a visual aid in the form of a red dot following the outline of a blue square, which moved synchronously to the audio instructions and increased/decreased in diameter during the inspiration/expiration phases.

### Control Condition

Everyday life was used as a control for the intervention periods in which the participants performed an anti-stress intervention. Participants were specifically instructed not to perform the anti-stress intervention during this time. PROs were documented in the same way as during the intervention period.

### Outcomes

Two primary outcomes were documented on every day by the participants within the StudyU App:

– “Overall, how stressful was your day?”

– “Which level of stress do you expect for tomorrow?”

Each item was answered on a visual analog scale from 1 (“not stressed at all”) to 10 (“extremely stressed out”). Additionally, participants were asked whether they performed the anti-stress exercise on a given day. All three items were available to be answered by the participants after 4 p.m. on every day and participants were reminded to document their PRO by push notification to their mobile phone.

### Effect modifiers and additional variables

Participants were asked to fill out an electronic baseline questionnaire hosted on REDCap (Research Electronic Data Capture)^68^. As part of this questionnaire, information on the participants’ demographics (e.g., age, sex, state of residence, relationship status, children in household), work (e.g., place of work, workhours per contract, average overtime), and lifestyle (e.g., alcohol, smoking, exercise) was collected., Additionally, validated German versions of established psychological questionnaires were assessed which included the Copenhagen Psychosocial Questionnaire (COPSOQ, work-life conflicts) ^69,70^, Copenhagen Burnout Inventory (Personal Burnout)^69^, Effort-Reward Imbalance Questionnaire (ERI)^71,72^, ERI Overcommitment^71,72^, Cohen’s Perceived Stress Scale^73^, Satisfaction with Life Scale (SWLS)^74^, and the System Usability Scale (SUS). The psychological scores were calculated as mean point scores, as suggested by the questionnaire authors. If more than 50% of the items per questionnaire were missing, no final score was calculated.

### Randomization

An alternating sequence of intervention (A) and control (B) periods was randomly assigned to each participant. Each sequence contained two pairs of alternating intervention and control periods resulting in two possible sequences: ABAB and BABA. Participants were randomly assigned to an individual linkage codes at the end of the baseline questionnaire. In the StudyU App each potential linkage code was filed prior to study start and alternately assigned to one of the two available randomization sequences (ABAB or BABA).

### Protocol Adherence

In addition to the daily assessment of the PROs, participants were asked on each day whether they did perform their anti-stress intervention. Protocol adherence was high, with 91.9% of all documented PROs aligning with the pre-specified protocol. Specifically, among participants evaluating Mindfulness Breathing, 244/246=99.2% (control phase) and 176/215=81.7% (intervention phase) of PROs were documented following correct protocol adherence. Among participants performing Box Breathing, 259/286=91.0% (control phase) and 317/340=93.2% (intervention phase) of all PROs aligned with the respective protocol phase.

### Follow-up

On October 1^st^ and 2^nd^, 2024, three months after completing the collection of data from the individual N-of-1 trials, an email with an invitation to fill out a follow-up questionnaire was sent out to all participants. On average, the follow-up questionnaire was filled out 18.0 weeks after completing the N-of-1 trial, and the individual follow-up time ranged between 13.9 and 20.7 weeks. This outcome was anticipated, as the data for the N-of-1 trials was collected over a 2.5-month period but all invitations to participate in the follow-up questionnaire were sent on the same date because, for anonymity reasons, the exact completion date of each individual trial was not known. Furthermore, the time between receiving the invitation and filling out the follow-up questionnaire differed between participants which added variance in the follow-up time.

Longitudinal data linkage was done using a seven-digit Self-Generated Identification Code (SGIC) derived from specifically designed questions at the beginning of the baseline and follow-up questionnaire:

– Please enter the first and second letter of your mother’s first name.

– Please enter the first and second letter of your place of birth.

– Please indicate your sex as stated in your birth certificate.

– Please enter the number of your older (not younger!) siblings.

– Please enter the last digit of your parent’s house number.

– Please enter the last digit of your parent’s postal code.

Overall, 39 participants filled out the follow-up questionnaire. Exact matching was possible for 27 cases (69.2%). Fuzzy matching allowing a one-digit difference between SGICs using R’s stringdist_join function (fuzzyjoin package) resulted in four additional matches which were subsequently confirmed using demographic information provided at both time points. Therefore, successful longitudinal matching was possible in 31/39=79.5% of all cases. Of the matched cases, one (Mindfulness Breathing) and two participants (Box Breathing) filled out baseline and follow-up questionnaires but did not participate in their N-of-1 trial and were therefore excluded from the analyses. Additionally, one participant from the Box Breathing group filled out the follow-up questionnaire but did not provide answers to the questions reported in Table 3.

### Data management

All data was collected anonymously, i.e. no connection of any of the collected data with identifying information was possible at any point in time. Data was collected electronically using two platforms. The electronic baseline and follow-up surveys were conducted using REDCap^68^. The anonymous data collected as part of the questionnaire is stored on a protected server at the Charité – Universitätsmedizin Berlin, Berlin, Germany. Daily PROs were assessed through the StudyU App, a mobile application installed on each participant’s smartphone^67^. The data collected through the StudyU App is stored on a secure backend host on servers located at the Hasso Plattner Institute in Potsdam, Germany. Data collected from the baseline questionnaire and data recorded with the StudyU App was linked using the individual linkage code given to each participant at the end of the baseline questionnaire and needed to access their N-of-1 trial in the StudyU App.

### Sample Size

The sample size was calculated using the approach described by Yang and colleagues^75^. Based on results from the literature ^76–78^, we assumed a homogenous residual standard error of 2.41, an autocorrelation of PROs of 0.8, and a reduction of the daily stress level by a standardized mean difference of at least 0.3. The level of statistical significance was set at α=0.05. A statistical power of 80% was reached when at least six participants per sequence reported at least four PROs per 1-week phase during a four-week trial period (per intervention: n=2*6=12). To account for a 30% dropout rate, n=34 participants were determined to be the required sample size for recruitment^59^.

### Statistical methods

Descriptive statistics of the study population are presented as mean, standard deviation, minimum, and maximum or number of observations and percentages in Table 1.

Bayesian multilevel models employing the Markov Chain Monte Carlo (MCMC) method are used to estimate the posterior distribution of the average effect of each of the two investigated interventions on the two primary outcomes: daily level of stress and daily level of stress expected for the following day. In the sampling procedure, we used two chains, 5000 burn-in steps, and 10,000 iterations. In the Bayesian analyses, a first-order autoregressive error structure (AR1) was used. Bayesian models allow a probabilistic description of the data and can be used to derive the probability of reaching a specified clinically relevant effect. Here, a clinically relevant effect was defined as a reduction of at least 0.5 points on the 0 to 10 stress scale. As the individual burden to perform the intervention is low, we defined participants with a probability of at least 70% for reaching the clinically relevant effect as responders. The one-sided cumulative probabilities of reaching effect sizes below zero and below 0.5 points were calculated.

An intention-to-treat (ITT) analysis was performed. Therefore, all available data points were included in the analyses independent of the number of reported outcomes or if the trial was completed. The role of potential effect measure modifiers was exploratively analyzed in subgroup analyses. All statistical analyses were conducted using the R software package version 4.4.1^79^. Bayesian models were calculated using the brms-package. Figures were drawn with the ggplot2 package.

### Ethics and Declarations Ethics

All participants provided informed consent. The trial was conducted in accordance with the Declaration of Helsinki and approved by the Ethics Committee of the Charité – Universitätsmedizin Berlin - approval number EA4/260/23. It is registered at ClinicalTrials.gov under trial ID NCT06368791 (first registered on April 11, 2024).

## Acknowledgments

No financial support was received for the research, authorship, and/or publication of this article. The authors would like to thank Ellen Zitzmann for her valuable input and help in developing the mindfulness-based breathing exercise and recording the intervention audio files.

## Data availability

The study setup, all information on protocol of the conducted N-of-1 as well as all data collected as part of the N-of-1 trials is publicly available through the StudyU data repository at https://designer.studyu.health (Mindfulness Breathing: “ASIP-Studie Anti-Stress Übung”, Box Breathing: “ASIP-Studie: Box-Atmung”). All data used to produce the results shown in this manuscript (including the data from the baseline and follow-up questionnaires collected through REDCap) is published at https://github.com/HIAlab/ASIP.

## Code availability

Statistical code used to produce all statistical analyses and figures is available at https://github.com/HIAlab/ASIP

## Author contributions

VMV: Conceptualization, Methodology, Formal analysis, Investigation, Data Curation, Writing – original draft, Writing – review & editing, Visualization, Project administration. TK: Conceptualization, Resources, Supervision, Writing – review & editing. SK: Conceptualization, Methodology, Resources, Supervision, Writing – review & editing.

## Supplementary Figures

**Supplementary Figure 1:**
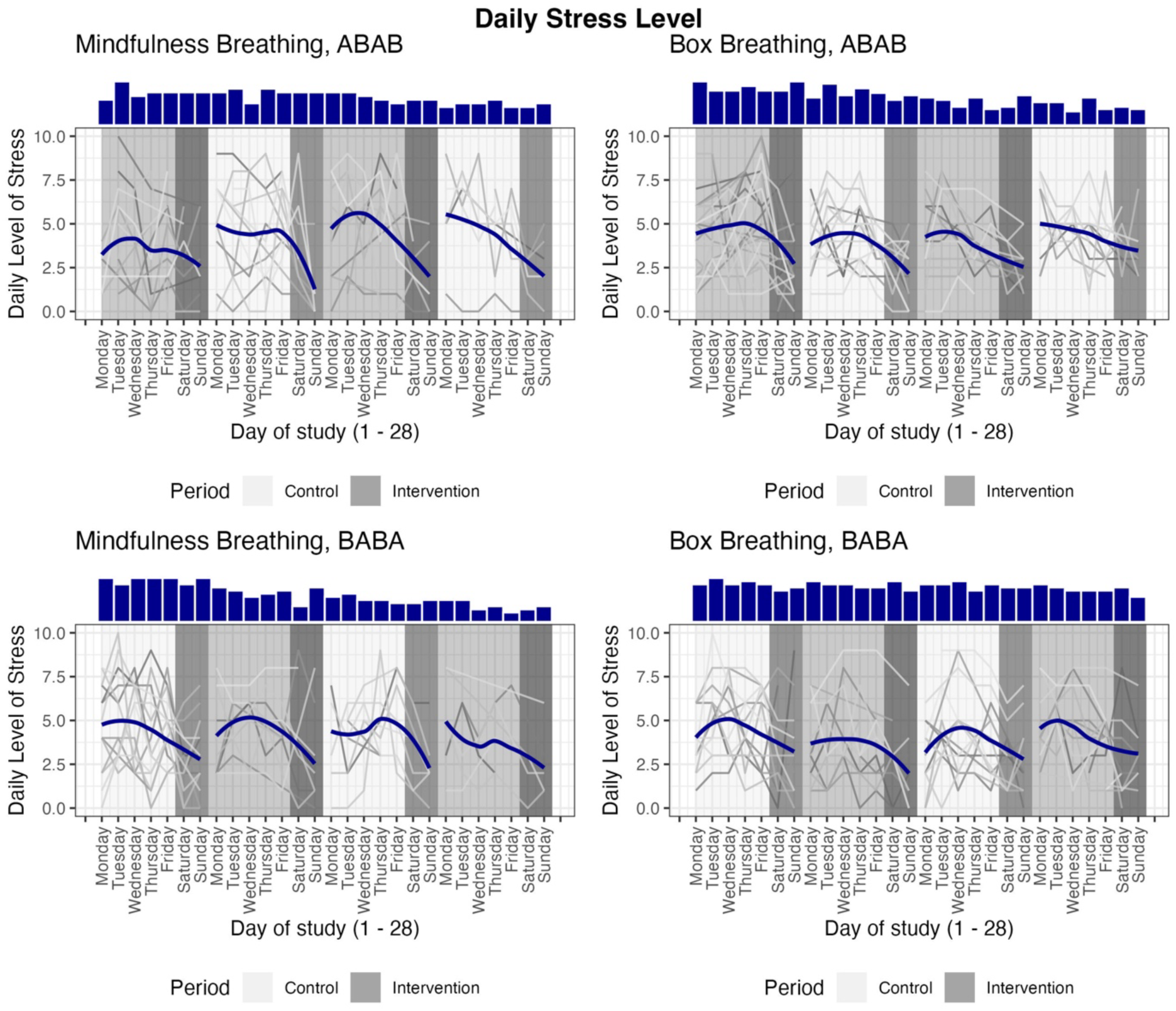
Individual line plots display the daily stress levels of each participant, stratified by their preferred intervention (Mindfulness Breathing or Box Breathing) and the randomly assigned trial sequence (A = intervention period, B: = control period). The blue line was fitted to all data points collected to allow interpretation of overall trends. The marginal histograms report the number of observations per day of study. To facilitate comparison, the data for each participant has been restructured to align on a common timeline. As a result, two consecutive data points may not correspond to consecutive days of data collection for each participant and the individual calendry day of study participation at one specific day on the x-axis differs between participants.

**Supplementary Figure 2:**
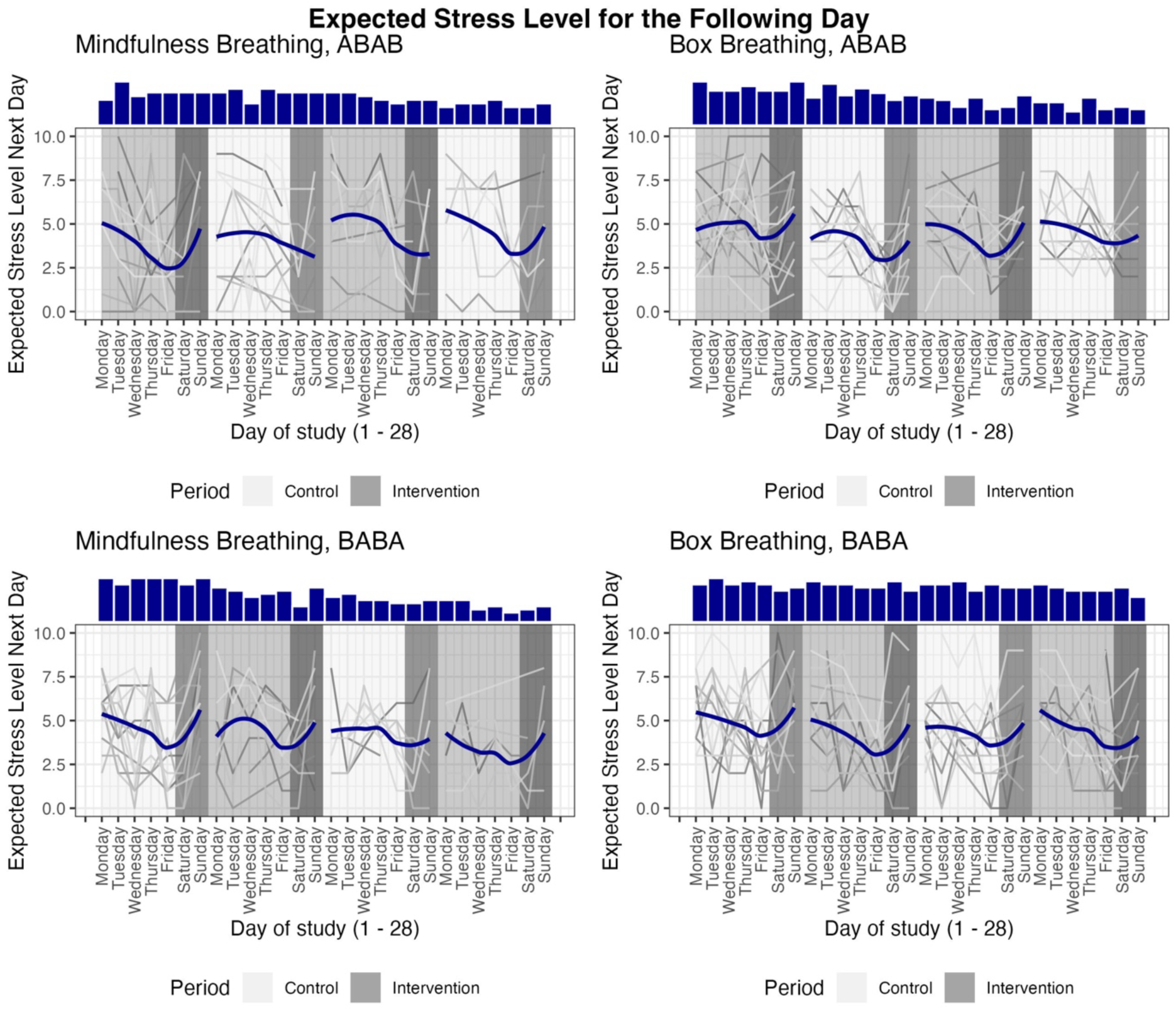
Individual line plots display the stress levels expected for the next day of each participant, stratified by their preferred intervention (Mindfulness Breathing or Box Breathing) and the randomly assigned trial sequence (A: intervention period, B: control period). The blue line was fitted to all data points collected to allow interpretation of overall trends. The marginal histograms report the number of observations per day of study. To facilitate comparison, the data for each participant has been restructured to align on a common timeline. As a result, two consecutive data points may not correspond to consecutive days of data collection for each participant and the individual calendry day of study participation at one specific day on the x-axis differs between participants.

## Supplementary Tables

**Supplementary Table 1:**
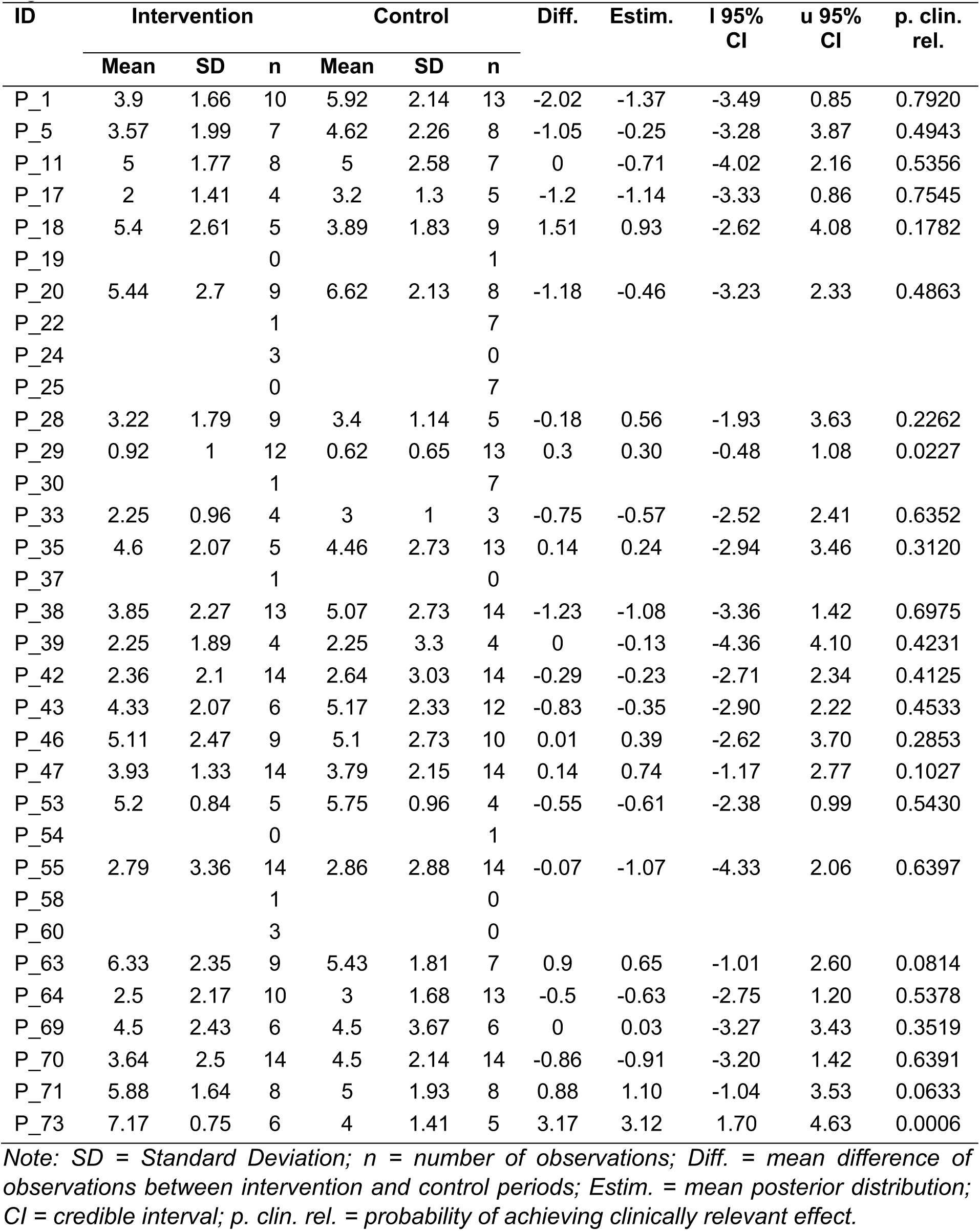
Individual level average intervention effect of performing Mindfulness Breathing on the daily stress level. Bayesian models using an autoregressive error structure (AR1) are used to estimate the mean posterior distribution and 95% credible interval. Additionally, the probability to reach a clinically relevant stress reduction of at least 0.5 points is given.

| ID | Intervention |  |  | Control |  |  | Diff. | Estim. | l 95% CI | u 95% CI | p. clin. rel. |
| --- | --- | --- | --- | --- | --- | --- | --- | --- | --- | --- | --- |
|  | Mean | SD | n | Mean | SD | n |  |  |  |  |  |
| P_1 | 3.9 | 1.66 | 10 | 5.92 | 2.14 | 13 | -2.02 | -1.37 | -3.49 | 0.85 | 0.7920 |
| P_5 | 3.57 | 1.99 | 7 | 4.62 | 2.26 | 8 | -1.05 | -0.25 | -3.28 | 3.87 | 0.4943 |
| P_11 | 5 | 1.77 | 8 | 5 | 2.58 | 7 | 0 | -0.71 | -4.02 | 2.16 | 0.5356 |
| P_17 | 2 | 1.41 | 4 | 3.2 | 1.3 | 5 | -1.2 | -1.14 | -3.33 | 0.86 | 0.7545 |
| P_18 | 5.4 | 2.61 | 5 | 3.89 | 1.83 | 9 | 1.51 | 0.93 | -2.62 | 4.08 | 0.1782 |
| P_19 |  |  | 0 |  |  | 1 |  |  |  |  |  |
| P_20 | 5.44 | 2.7 | 9 | 6.62 | 2.13 | 8 | -1.18 | -0.46 | -3.23 | 2.33 | 0.4863 |
| P_22 |  |  | 1 |  |  | 7 |  |  |  |  |  |
| P_24 |  |  | 3 |  |  | 0 |  |  |  |  |  |
| P_25 |  |  | 0 |  |  | 7 |  |  |  |  |  |
| P_28 | 3.22 | 1.79 | 9 | 3.4 | 1.14 | 5 | -0.18 | 0.56 | -1.93 | 3.63 | 0.2262 |
| P_29 | 0.92 | 1 | 12 | 0.62 | 0.65 | 13 | 0.3 | 0.30 | -0.48 | 1.08 | 0.0227 |
| P_30 |  |  | 1 |  |  | 7 |  |  |  |  |  |
| P_33 | 2.25 | 0.96 | 4 | 3 | 1 | 3 | -0.75 | -0.57 | -2.52 | 2.41 | 0.6352 |
| P_35 | 4.6 | 2.07 | 5 | 4.46 | 2.73 | 13 | 0.14 | 0.24 | -2.94 | 3.46 | 0.3120 |
| P_37 |  |  | 1 |  |  | 0 |  |  |  |  |  |
| P_38 | 3.85 | 2.27 | 13 | 5.07 | 2.73 | 14 | -1.23 | -1.08 | -3.36 | 1.42 | 0.6975 |
| P_39 | 2.25 | 1.89 | 4 | 2.25 | 3.3 | 4 | 0 | -0.13 | -4.36 | 4.10 | 0.4231 |
| P_42 | 2.36 | 2.1 | 14 | 2.64 | 3.03 | 14 | -0.29 | -0.23 | -2.71 | 2.34 | 0.4125 |
| P_43 | 4.33 | 2.07 | 6 | 5.17 | 2.33 | 12 | -0.83 | -0.35 | -2.90 | 2.22 | 0.4533 |
| P_46 | 5.11 | 2.47 | 9 | 5.1 | 2.73 | 10 | 0.01 | 0.39 | -2.62 | 3.70 | 0.2853 |
| P_47 | 3.93 | 1.33 | 14 | 3.79 | 2.15 | 14 | 0.14 | 0.74 | -1.17 | 2.77 | 0.1027 |
| P_53 | 5.2 | 0.84 | 5 | 5.75 | 0.96 | 4 | -0.55 | -0.61 | -2.38 | 0.99 | 0.5430 |
| P_54 |  |  | 0 |  |  | 1 |  |  |  |  |  |
| P_55 | 2.79 | 3.36 | 14 | 2.86 | 2.88 | 14 | -0.07 | -1.07 | -4.33 | 2.06 | 0.6397 |
| P_58 |  |  | 1 |  |  | 0 |  |  |  |  |  |
| P_60 |  |  | 3 |  |  | 0 |  |  |  |  |  |
| P_63 | 6.33 | 2.35 | 9 | 5.43 | 1.81 | 7 | 0.9 | 0.65 | -1.01 | 2.60 | 0.0814 |
| P_64 | 2.5 | 2.17 | 10 | 3 | 1.68 | 13 | -0.5 | -0.63 | -2.75 | 1.20 | 0.5378 |
| P_69 | 4.5 | 2.43 | 6 | 4.5 | 3.67 | 6 | 0 | 0.03 | -3.27 | 3.43 | 0.3519 |
| P_70 | 3.64 | 2.5 | 14 | 4.5 | 2.14 | 14 | -0.86 | -0.91 | -3.20 | 1.42 | 0.6391 |
| P_71 | 5.88 | 1.64 | 8 | 5 | 1.93 | 8 | 0.88 | 1.10 | -1.04 | 3.53 | 0.0633 |
| P_73 | 7.17 | 0.75 | 6 | 4 | 1.41 | 5 | 3.17 | 3.12 | 1.70 | 4.63 | 0.0006 |

Note: SD = Standard Deviation; n = number of observations; Diff. = mean difference of observations between intervention and control periods; Estim. = mean posterior distribution; CI = credible interval; p. clin. rel. = probability of achieving clinically relevant effect.

**Supplementary Table 2:** Individual level average intervention effect of performing the Box Breathing on the daily stress level. Bayesian models using an autoregressive error structure (AR1) are used to estimate the mean posterior distribution and 95% credible interval. Additionally, the probability to reach a clinically relevant stress reduction of at least 0.5 points is given.

| ID | Intervention |  |  | Control |  |  | Diff. | Estim. | l 95% CI | u 95% CI | p. clin. rel. |
| --- | --- | --- | --- | --- | --- | --- | --- | --- | --- | --- | --- |
|  | Mean | SD | n | Mean | SD | n |  |  |  |  |  |
| P_2 | 3.36 | 1.95 | 14 | 3.5 | 2.21 | 14 | -0.14 | -0.17 | -2.23 | 1.82 | 0.3627 |
| P_3 | 1.6 | 1.65 | 10 | 2.62 | 1.8 | 13 | -1.02 | -1.04 | -2.38 | 0.34 | 0.8055 |
| P_4 | 4.09 | 1.81 | 11 | 3.67 | 1.87 | 12 | 0.42 | 0.54 | -1.48 | 2.59 | 0.1439 |
| P_6 | 4 | 2.37 | 11 | 3.9 | 1.66 | 10 | 0.1 | -0.17 | -2.93 | 1.96 | 0.3507 |
| P_7 | 5.08 | 1.93 | 13 | 3.25 | 1.6 | 12 | 1.83 | 1.50 | -0.56 | 3.39 | 0.0284 |
| P_8 | 5.12 | 1.96 | 8 | 2.71 | 0.76 | 7 | 2.41 | 2.45 | 0.49 | 4.47 | 0.0034 |
| P_9 | 5.6 | 3.05 | 5 | 4.67 | 1.53 | 3 | 0.93 | 0.07 | -6.42 | 5.04 | 0.3728 |
| P_10 |  |  | 4 |  |  | 1 |  |  |  |  |  |
| P_12 | 2.43 | 1.81 | 7 | 4 | 2.19 | 6 | -1.57 | -2.18 | -5.29 | 0.75 | 0.8831 |
| P_13 |  |  | 2 |  |  | 0 |  |  |  |  |  |
| P_14 | 4 | 2 | 9 | 4.38 | 1.41 | 8 | -0.38 | -0.08 | -2.11 | 2.44 | 0.3773 |
| P_15 |  |  | 2 |  |  | 1 |  |  |  |  |  |
| P_16 | 2.56 | 1.51 | 9 | 5.67 | 2.8 | 6 | -3.11 | -2.97 | -5.51 | 0.01 | 0.9546 |
| P_21 | 4.62 | 2.45 | 8 | 3.14 | 2.19 | 7 | 1.48 | 1.68 | -1.26 | 4.73 | 0.065 |
| P_23 |  |  | 2 |  |  | 0 |  |  |  |  |  |
| P_26 | 4.45 | 0.69 | 11 | 4 | 1.86 | 12 | 0.45 | 0.57 | -1.04 | 2.19 | 0.0894 |
| P_27 |  |  | 3 |  |  | 0 |  |  |  |  |  |
| P_31 | 6 | 1.41 | 2 | 6.17 | 1.17 | 6 | -0.17 | 0.12 | -2.43 | 2.61 | 0.2911 |
| P_32 | 4.14 | 1.68 | 7 | 4.6 | 2.07 | 5 | -0.46 | -0.30 | -3.44 | 1.91 | 0.3815 |
| P_34 |  |  | 6 |  |  | 1 |  |  |  |  |  |
| P_36 | 2.27 | 0.9 | 11 | 3.67 | 0.98 | 12 | -1.39 | -1.34 | -2.20 | -0.61 | 0.9854 |
| P_40 | 4 | 1.07 | 8 | 5 | 0.93 | 8 | -1 | -1.00 | -2.28 | 0.29 | 0.7938 |
| P_41 | 2.31 | 1.32 | 13 | 3.31 | 2.87 | 13 | -1 | -0.39 | -2.71 | 2.37 | 0.487 |
| P_44 |  |  | 0 |  |  | 1 |  |  |  |  |  |
| P_45 |  |  | 3 |  |  | 0 |  |  |  |  |  |
| P_48 | 3.54 | 2.18 | 13 | 2.45 | 1.97 | 11 | 1.08 | 0.82 | -1.52 | 2.99 | 0.119 |
| P_49 |  |  | 3 |  |  | 0 |  |  |  |  |  |
| P_50 | 4.08 | 2.81 | 12 | 5.36 | 2.5 | 14 | -1.27 | -1.29 | -3.74 | 0.88 | 0.7644 |
| P_51 |  |  | 4 |  |  | 1 |  |  |  |  |  |
| P_52 | 3 | 1.47 | 14 | 3.79 | 1.58 | 14 | -0.79 | -0.74 | -2.19 | 0.77 | 0.633 |
| P_56 | 2.8 | 2.28 | 5 | 4.5 | 2.66 | 6 | -1.7 | -1.74 | -5.88 | 1.71 | 0.7616 |
| P_57 |  |  | 0 |  |  | 4 |  |  |  |  |  |
| P_59 | 3.38 | 0.92 | 8 | 4.1 | 0.88 | 10 | -0.72 | -0.60 | -1.67 | 0.56 | 0.586 |
| P_61 | 2.57 | 1.27 | 7 | 2.36 | 1.12 | 11 | 0.21 | 0.73 | -0.75 | 2.26 | 0.0489 |
| P_62 | 4.62 | 1.61 | 13 | 4.6 | 2.41 | 10 | 0.02 | -0.16 | -2.40 | 2.07 | 0.369 |
| P_65 | 6.08 | 1.38 | 12 | 5.31 | 1.6 | 13 | 0.78 | 0.71 | -0.82 | 2.15 | 0.055 |
| P_66 | 4 | 1.31 | 8 | 3.8 | 0.92 | 10 | 0.2 | 0.25 | -1.11 | 1.66 | 0.1243 |
| P_67 | 1.57 | 0.79 | 7 | 1 | 1 | 7 | 0.57 | 0.50 | -1.06 | 1.65 | 0.0705 |
| P_68 | 5.75 | 1.82 | 12 | 5.5 | 1.61 | 14 | 0.25 | 0.24 | -1.16 | 1.68 | 0.1421 |
| P_72 | 5 | 2.06 | 9 | 4.89 | 0.78 | 9 | 0.11 | 0.07 | -1.80 | 1.87 | 0.2494 |
| P_74 | 3.14 | 1.83 | 14 | 3.77 | 1.48 | 13 | -0.63 | -0.62 | -2.02 | 0.71 | 0.5656 |
| P_75 | 7.33 | 1.32 | 9 | 6.92 | 2.19 | 12 | 0.42 | 0.33 | -1.91 | 2.51 | 0.2125 |
| P_76 | 2.82 | 2.75 | 11 | 2.6 | 1.67 | 5 | 0.22 | -1.03 | -4.52 | 2.49 | 0.6169 |

Note: SD = Standard Deviation; n = number of observations; Diff. = mean difference of observations between intervention and control periods; Estim. = mean posterior distribution; CI = credible interval; p. clin. rel. = probability of achieving clinically relevant effect.

**Supplementary Table 3:** Individual-level average intervention effect of performing the Mindfulness Breathing on the stress level expected for the next day. Bayesian models using an autoregressive error structure (AR1) are used to estimate the mean posterior distribution and 95% credible interval. Additionally, the probability to reach a clinically relevant stress reduction of at least 0.5 points is given.

| ID | Intervention |  |  | Control |  |  | Diff. | Esti<br>m. | l<br>95%<br>CI | u<br>95%<br>CI | p. clin.<br>rel. |
| --- | --- | --- | --- | --- | --- | --- | --- | --- | --- | --- | --- |
|  | Mea<br>n | SD | n | Mea<br>n | SD | n |  |  | Mea<br>n | SD | n |
| P_1 | 3.9 | 1.66 | 10 | 5.92 | 2.14 | 13 | -2.02 | -1.18 | -3.50 | 1.27 | 0.7283 |
| P_5 | 3.57 | 1.99 | 7 | 4.62 | 2.26 | 8 | -1.05 | 0.00 | -3.15 | 3.61 | 0.3944 |
| P_11 | 5 | 1.77 | 8 | 5 | 2.58 | 7 | 0 | -0.31 | -3.54 | 2.13 | 0.3944 |
| P_17 | 2 | 1.41 | 4 | 3.2 | 1.3 | 5 | -1.2 | -1.18 | -3.06 | 0.60 | 0.8039 |
| P_18 | 5.4 | 2.61 | 5 | 3.89 | 1.83 | 9 | 1.51 | -0.39 | -2.62 | 1.91 | 0.4554 |
| P_19 |  |  | 0 |  |  | 1 |  |  |  |  |  |
| P_20 | 5.44 | 2.7 | 9 | 6.62 | 2.13 | 8 | -1.18 | -0.86 | -2.70 | 0.78 | 0.6697 |
| P_22 |  |  | 1 |  |  | 7 |  |  |  |  |  |
| P_24 |  |  | 3 |  |  | 0 |  |  |  |  |  |
| P_25 |  |  | 0 |  |  | 7 |  |  |  |  |  |
| P_28 | 3.22 | 1.79 | 9 | 3.4 | 1.14 | 5 | -0.18 | -0.01 | -2.30 | 2.29 | 0.3259 |
| P_29 | 0.92 | 1 | 12 | 0.62 | 0.65 | 13 | 0.3 | 0.20 | -1.15 | 1.47 | 0.1384 |
| P_30 |  |  | 1 |  |  | 7 |  |  |  |  |  |
| P_33 | 2.25 | 0.96 | 4 | 3 | 1 | 3 | -0.75 | 0.71 | -3.00 | 4.03 | 0.2133 |
| P_35 | 4.6 | 2.07 | 5 | 4.46 | 2.73 | 13 | 0.14 | 0.29 | -1.98 | 2.58 | 0.2404 |
| P_37 |  |  | 1 |  |  | 0 |  |  |  |  |  |
| P_38 | 3.85 | 2.27 | 13 | 5.07 | 2.73 | 14 | -1.23 | -0.16 | -2.96 | 2.72 | 0.4024 |
| P_39 | 2.25 | 1.89 | 4 | 2.25 | 3.3 | 4 | 0 | 0.25 | -1.44 | 1.78 | 0.1438 |
| P_42 | 2.36 | 2.1 | 14 | 2.64 | 3.03 | 14 | -0.29 | -0.31 | -2.84 | 3.21 | 0.4864 |
| P_43 | 4.33 | 2.07 | 6 | 5.17 | 2.33 | 12 | -0.83 | 0.50 | -1.67 | 2.92 | 0.1761 |
| P_46 | 5.11 | 2.47 | 9 | 5.1 | 2.73 | 10 | 0.01 | -0.77 | -2.71 | 1.21 | 0.6195 |
| P_47 | 3.93 | 1.33 | 14 | 3.79 | 2.15 | 14 | 0.14 | -0.57 | -2.39 | 1.24 | 0.5338 |
| P_53 | 5.2 | 0.84 | 5 | 5.75 | 0.96 | 4 | -0.55 | 0.99 | -0.99 | 2.62 | 0.059 |
| P_54 |  |  | 0 |  |  | 1 |  |  |  |  |  |
| P_55 | 2.79 | 3.36 | 14 | 2.86 | 2.88 | 14 | -0.07 | 2.29 | -0.80 | 5.98 | 0.0386 |
| P_58 |  |  | 1 |  |  | 0 |  |  |  |  |  |
| P_60 |  |  | 3 |  |  | 0 |  |  |  |  |  |
| P_63 | 6.33 | 2.35 | 9 | 5.43 | 1.81 | 7 | 0.9 | 0.25 | -1.75 | 1.97 | 0.174 |
| P_64 | 2.5 | 2.17 | 10 | 3 | 1.68 | 13 | -0.5 | -0.67 | -2.63 | 1.35 | 0.5732 |
| P_69 | 4.5 | 2.43 | 6 | 4.5 | 3.67 | 6 | 0 | -1.67 | -7.94 | 2.06 | 0.6651 |
| P_70 | 3.64 | 2.5 | 14 | 4.5 | 2.14 | 14 | -0.86 | -0.64 | -3.29 | 2.28 | 0.5542 |
| P_71 | 5.88 | 1.64 | 8 | 5 | 1.93 | 8 | 0.88 | 0.10 | -2.27 | 2.42 | 0.2829 |
| P_73 | 7.17 | 0.75 | 6 | 4 | 1.41 | 5 | 3.17 | 1.75 | -0.51 | 3.87 | 0.0255 |

Note: SD = Standard Deviation; n = number of observations; Diff. = mean difference of observations between intervention and control periods; Estim. = mean posterior distribution; CI = credible interval; p. clin. rel. = probability of achieving clinically relevant effect.

**Supplementary Table 4:**
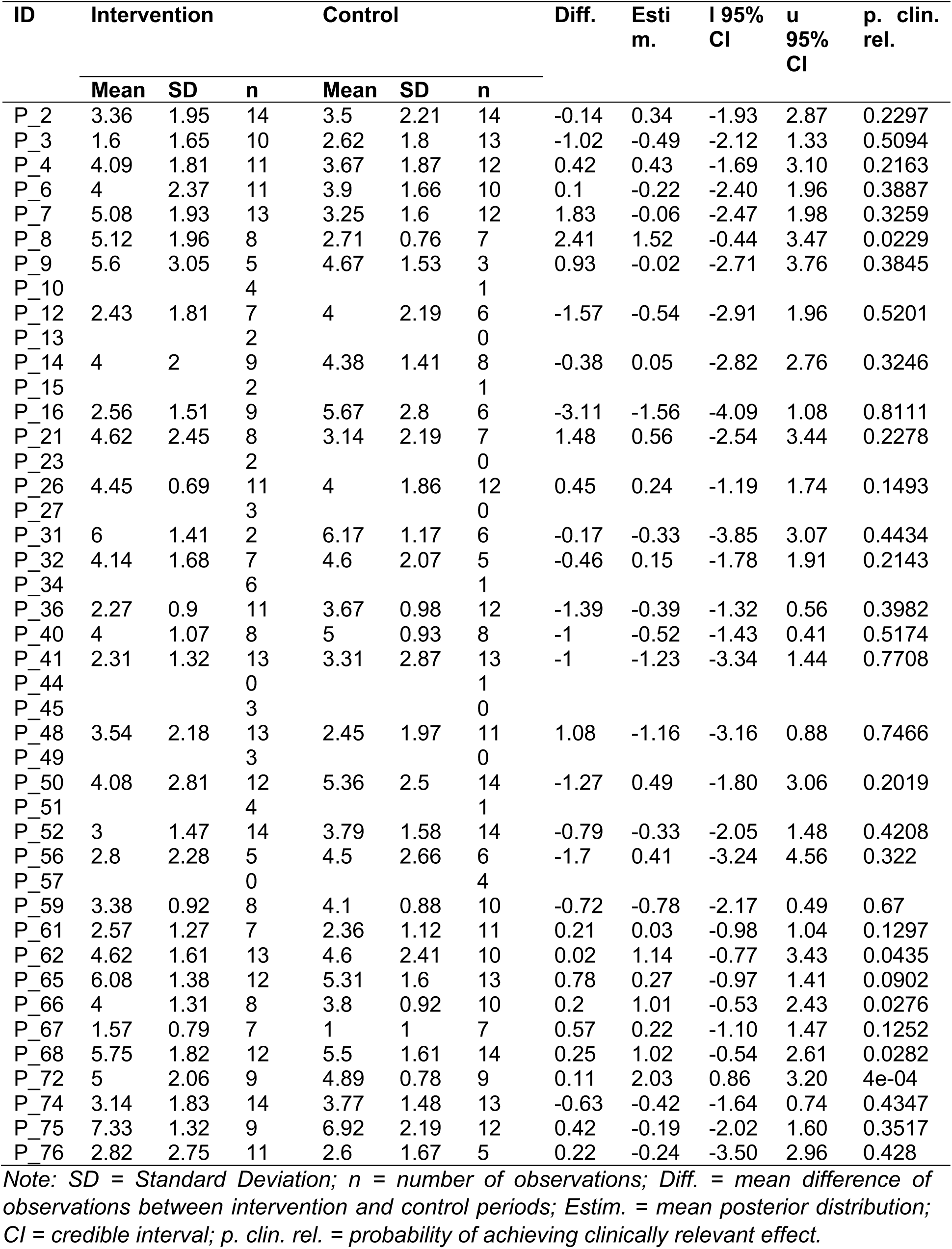
Individual-level average intervention effect of performing the Box Breathing on the stress level expected for the next day. Bayesian models using an autoregressive error structure (AR1) are used to estimate the mean posterior distribution and 95% credible interval. Additionally, the probability to reach a clinically relevant stress reduction of at least 0.5 points is given.

| ID | Intervention |  |  | Control |  |  | Diff. | Esti<br>m. | l 95%<br>CI | u 95%<br>CI | p. clin.<br>rel. |
| --- | --- | --- | --- | --- | --- | --- | --- | --- | --- | --- | --- |
|  | Mean | SD | n | Mean | SD | n |  |  |  |  |  |
| P_2 | 3.36 | 1.95 | 14 | 3.5 | 2.21 | 14 | -0.14 | 0.34 | -1.93 | 2.87 | 0.2297 |
| P_3 | 1.6 | 1.65 | 10 | 2.62 | 1.8 | 13 | -1.02 | -0.49 | -2.12 | 1.33 | 0.5094 |
| P_4 | 4.09 | 1.81 | 11 | 3.67 | 1.87 | 12 | 0.42 | 0.43 | -1.69 | 3.10 | 0.2163 |
| P_6 | 4 | 2.37 | 11 | 3.9 | 1.66 | 10 | 0.1 | -0.22 | -2.40 | 1.96 | 0.3887 |
| P_7 | 5.08 | 1.93 | 13 | 3.25 | 1.6 | 12 | 1.83 | -0.06 | -2.47 | 1.98 | 0.3259 |
| P_8 | 5.12 | 1.96 | 8 | 2.71 | 0.76 | 7 | 2.41 | 1.52 | -0.44 | 3.47 | 0.0229 |
| P_9 | 5.6 | 3.05 | 5 | 4.67 | 1.53 | 3 | 0.93 | -0.02 | -2.71 | 3.76 | 0.3845 |
| P_10 |  |  | 4 |  |  | 1 |  |  |  |  |  |
| P_12 | 2.43 | 1.81 | 7 | 4 | 2.19 | 6 | -1.57 | -0.54 | -2.91 | 1.96 | 0.5201 |
| P_13 |  |  | 2 |  |  | 0 |  |  |  |  |  |
| P_14 | 4 | 2 | 9 | 4.38 | 1.41 | 8 | -0.38 | 0.05 | -2.82 | 2.76 | 0.3246 |
| P_15 |  |  | 2 |  |  | 1 |  |  |  |  |  |
| P_16 | 2.56 | 1.51 | 9 | 5.67 | 2.8 | 6 | -3.11 | -1.56 | -4.09 | 1.08 | 0.8111 |
| P_21 | 4.62 | 2.45 | 8 | 3.14 | 2.19 | 7 | 1.48 | 0.56 | -2.54 | 3.44 | 0.2278 |
| P_23 |  |  | 2 |  |  | 0 |  |  |  |  |  |
| P_26 | 4.45 | 0.69 | 11 | 4 | 1.86 | 12 | 0.45 | 0.24 | -1.19 | 1.74 | 0.1493 |
| P_27 |  |  | 3 |  |  | 0 |  |  |  |  |  |
| P_31 | 6 | 1.41 | 2 | 6.17 | 1.17 | 6 | -0.17 | -0.33 | -3.85 | 3.07 | 0.4434 |
| P_32 | 4.14 | 1.68 | 7 | 4.6 | 2.07 | 5 | -0.46 | 0.15 | -1.78 | 1.91 | 0.2143 |
| P_34 |  |  | 6 |  |  | 1 |  |  |  |  |  |
| P_36 | 2.27 | 0.9 | 11 | 3.67 | 0.98 | 12 | -1.39 | -0.39 | -1.32 | 0.56 | 0.3982 |
| P_40 | 4 | 1.07 | 8 | 5 | 0.93 | 8 | -1 | -0.52 | -1.43 | 0.41 | 0.5174 |
| P_41 | 2.31 | 1.32 | 13 | 3.31 | 2.87 | 13 | -1 | -1.23 | -3.34 | 1.44 | 0.7708 |
| P_44 |  |  | 0 |  |  | 1 |  |  |  |  |  |
| P_45 |  |  | 3 |  |  | 0 |  |  |  |  |  |
| P_48 | 3.54 | 2.18 | 13 | 2.45 | 1.97 | 11 | 1.08 | -1.16 | -3.16 | 0.88 | 0.7466 |
| P_49 |  |  | 3 |  |  | 0 |  |  |  |  |  |
| P_50 | 4.08 | 2.81 | 12 | 5.36 | 2.5 | 14 | -1.27 | 0.49 | -1.80 | 3.06 | 0.2019 |
| P_51 |  |  | 4 |  |  | 1 |  |  |  |  |  |
| P_52 | 3 | 1.47 | 14 | 3.79 | 1.58 | 14 | -0.79 | -0.33 | -2.05 | 1.48 | 0.4208 |
| P_56 | 2.8 | 2.28 | 5 | 4.5 | 2.66 | 6 | -1.7 | 0.41 | -3.24 | 4.56 | 0.322 |
| P_57 |  |  | 0 |  |  | 4 |  |  |  |  |  |
| P_59 | 3.38 | 0.92 | 8 | 4.1 | 0.88 | 10 | -0.72 | -0.78 | -2.17 | 0.49 | 0.67 |
| P_61 | 2.57 | 1.27 | 7 | 2.36 | 1.12 | 11 | 0.21 | 0.03 | -0.98 | 1.04 | 0.1297 |
| P_62 | 4.62 | 1.61 | 13 | 4.6 | 2.41 | 10 | 0.02 | 1.14 | -0.77 | 3.43 | 0.0435 |
| P_65 | 6.08 | 1.38 | 12 | 5.31 | 1.6 | 13 | 0.78 | 0.27 | -0.97 | 1.41 | 0.0902 |
| P_66 | 4 | 1.31 | 8 | 3.8 | 0.92 | 10 | 0.2 | 1.01 | -0.53 | 2.43 | 0.0276 |
| P_67 | 1.57 | 0.79 | 7 | 1 | 1 | 7 | 0.57 | 0.22 | -1.10 | 1.47 | 0.1252 |
| P_68 | 5.75 | 1.82 | 12 | 5.5 | 1.61 | 14 | 0.25 | 1.02 | -0.54 | 2.61 | 0.0282 |
| P_72 | 5 | 2.06 | 9 | 4.89 | 0.78 | 9 | 0.11 | 2.03 | 0.86 | 3.20 | 4e-04 |
| P_74 | 3.14 | 1.83 | 14 | 3.77 | 1.48 | 13 | -0.63 | -0.42 | -1.64 | 0.74 | 0.4347 |
| P_75 | 7.33 | 1.32 | 9 | 6.92 | 2.19 | 12 | 0.42 | -0.19 | -2.02 | 1.60 | 0.3517 |
| P_76 | 2.82 | 2.75 | 11 | 2.6 | 1.67 | 5 | 0.22 | -0.24 | -3.50 | 2.96 | 0.428 |

Note: SD = Standard Deviation; n = number of observations; Diff. = mean difference of observations between intervention and control periods; Estim. = mean posterior distribution; CI = credible interval; p. clin. rel. = probability of achieving clinically relevant effect.

**Supplementary Table 5:** Sex-stratified mean posterior, 95% credible interval and posterior probability to reach a clinically relevant stress reduction of at least 0.5 points in 60 women and 17 men participating in the ASIP study. Additionally, sex-stratified mean, standard deviation, number of documented PROs and mean difference in outcome variables between intervention and control periods are shown.

| Outcome | Anti-Stress Exercise | Intervention Period |  |  | Control Period |  |  | Diff. | Estim. | Lower 95% CI | Upper 95% CI | p. clin. rel. |
| --- | --- | --- | --- | --- | --- | --- | --- | --- | --- | --- | --- | --- |
|  |  | Mean | SD | n | Mean | SD | n |  |  |  |  |  |
| Women |  |  |  |  |  |  |  |  |  |  |  |  |
| Daily Stress | Mindfulness Breathing | 3.94 | 2.54 | 199 | 3.89 | 2.6 | 214 | 0.05 | -0.03 | -0.50 | 0.45 | 0.025 |
|  | Box Breathing | 4.11 | 2.22 | 241 | 4.3 | 1.99 | 219 | -0.19 | -0.09 | -0.52 | 0.34 | 0.0298 |
| Stress Next Day | Mindfulness Breathing | 4.07 | 2.75 | 199 | 4.15 | 2.7 | 214 | -0.09 | -0.17 | -0.61 | 0.27 | 0.0724 |
|  | Box Breathing | 4.54 | 2.22 | 241 | 4.54 | 1.97 | 219 | 0 | 0.14 | -0.25 | 0.54 | 0.0005 |
| Men |  |  |  |  |  |  |  |  |  |  |  |  |
| Daily Stress | Mindfulness Breathing | 4.06 | 1.77 | 16 | 5.38 | 2.17 | 32 | -1.31 | -1.25 | -4.32 | 2.40 | 0.7493 |
|  | Box Breathing | 3.42 | 2.25 | 90 | 3.32 | 2.15 | 95 | 0.11 | 0.11 | -0.69 | 0.95 | 0.0666 |
| Stress Next Day | Mindfulness Breathing | 4.31 | 1.96 | 16 | 5.22 | 1.95 | 32 | -0.91 | -0.57 | -6.77 | 3.39 | 0.4837 |
|  | Box Breathing | 4.17 | 2.37 | 90 | 3.91 | 2.41 | 95 | 0.26 | 0.24 | -0.44 | 0.96 | 0.0179 |

Note: SD = Standard Deviation; n = number of observations; Diff. = Difference between average stress levels in intervention and control period; Estim. = mean posterior distribution; CI = credible interval; p. clin. rel. = probability of achieving clinically relevant effect.

**Supplementary Table 6:** Mean posterior, 95% credible interval and posterior probability to reach a clinically relevant stress reduction of at least 0.5 points stratified by baseline stress levels assessed by Cohen’s PSS. A median split was performed to compare participants below the 50^th^ percentile with participants above the 50^th^ percentile. Additionally, mean, standard deviation, number of documented PROs and mean difference in outcome variables between intervention and control periods are shown for the respective subgroups.

| Outcome | Anti-Stress Exercise | Intervention Period |  |  | Control Period |  |  | Diff. | Estim. | Lower 95% CI | Upper 95% CI | % clin. rel. |
| --- | --- | --- | --- | --- | --- | --- | --- | --- | --- | --- | --- | --- |
|  |  | Mean | SD | n | Mean | SD | n |  |  |  |  |  |
| Low Chronic Stress at Baseline |  |  |  |  |  |  |  |  |  |  |  |  |
| Daily Stress | Mindfulness Breathing | 3.75 | 2.29 | 96 | 3.79 | 2.64 | 120 | -0.04 | -0.01 | -0.71 | 0.71 | 0.0753 |
|  | Box Breathing | 3.30 | 2 | 174 | 3.43 | 2.04 | 152 | -0.14 | -0.32 | -0.97 | 0.31 | 0.269 |
| Stress Next Day | Mindfulness Breathing | 3.68 | 2.56 | 96 | 3.85 | 2.64 | 120 | -0.17 | -0.05 | -0.67 | 0.59 | 0.0704 |
|  | Box Breathing | 3.69 | 2.06 | 174 | 3.78 | 1.84 | 152 | -0.09 | -0.17 | -0.65 | 0.36 | 0.0877 |
| High Chronic Stress at Baseline |  |  |  |  |  |  |  |  |  |  |  |  |
| Daily Stress | Mindfulness Breathing | 4.11 | 2.64 | 119 | 4.37 | 2.52 | 126 | -0.26 | -0.27 | -0.95 | 0.40 | 0.2482 |
|  | Box Breathing | 4.58 | 2.29 | 166 | 4.52 | 1.97 | 170 | 0.05 | 0.13 | -0.35 | 0.64 | 0.0047 |
| Stress Next Day | Mindfulness Breathing | 4.41 | 2.78 | 119 | 4.71 | 2.57 | 126 | -0.3 | -0.29 | -0.97 | 0.38 | 0.2647 |
|  | Box Breathing | 5.21 | 2.22 | 166 | 4.85 | 2.25 | 170 | 0.36 | 0.38 | -0.06 | 0.83 | 0.0001 |

Note: SD = Standard Deviation; n = number of observations; Diff. = Difference between average stress levels in intervention and control period; Estim. = mean posterior distribution; CI = credible interval; p. clin. rel. = probability of achieving clinically relevant effect, PSS = Perceived Stress Scale.

**Supplementary Table 7:** Average intervention effect on the daily stress level and the level of stress expected for the following day of to the availability of an anti-stress intervention among the participants that chose the respective intervention. The dataset analyzed here contains all PROs documented by participants that chose to evaluate Mindfulness Breathing (intervention 1) or Box Breathing (intervention 2). Mean posterior, 95% credible interval and probability to reach a reduction of at least 0.5 points on the respective stress scales were estimated by Bayesian multilevel models. Mean, standard deviation and number of documented PROs in the analyzed dataset are displayed.

| Data and Outcome | Intervention Period |  |  | Control Period |  |  | Diff. | Estim. | Lower<br>95%<br>CI | Upper<br>95%<br>CI | p. clin. rel. |
| --- | --- | --- | --- | --- | --- | --- | --- | --- | --- | --- | --- |
|  | Mean | SD | n | Mean | SD | n |  |  |  |  |  |
| Across interventions (1 + 2), Daily Stress | 3.93 | 2.34 | 555 | 4.04 | 2.31 | 568 | -0.11 | -0.10 | -0.38 | 0.17 | 0.0026 |
| Across interventions (1 + 2), Stress Next Day | 4.30 | 2.45 | 555 | 4.32 | 2.36 | 568 | -0.02 | 0.00 | -0.24 | 0.24 | <0.0001 |

Note: SD = Standard Deviation; n = number of observations; Diff. = Difference between average stress levels in intervention and control period; Estim. = mean posterior distribution; CI = credible interval; p. clin. rel. = probability of achieving clinically relevant effect.

